# Treatment of Recurrent Glioblastoma by Chronic Convection-Enhanced Delivery of Topotecan

**DOI:** 10.1101/2021.12.04.21266935

**Authors:** Eleonora F. Spinazzi, Michael G. Argenziano, Pavan S. Upadhyayula, Matei A. Banu, Justin A. Neira, Dominique M.O. Higgins, Peter B. Wu, Brianna Pereira, Aayushi Mahajan, Nelson Humala, Osama Al-Dalahmah, Wenting Zhao, Akshay V. Save, Deborah M. Boyett, Tamara Marie, Julia L Furnari, Tejaswi D. Sudhakar, Sylwia A. Stopka, Michael S. Regan, Vanessa Catania, Laura Good, Meenu Behl, Sachin Jambawalikar, Akiva Mintz, Angela Lignelli, Nathalie Y.R. Agar, Peter A. Sims, Mary Welch, Andrew Lassman, Fabio Iwamoto, Randy S. D’Amico, Jack Grinband, Peter Canoll, Jeffrey N. Bruce

## Abstract

Glioblastoma, the most common primary brain malignancy, is invariably fatal. Systemic chemotherapy is ineffective mostly because of drug delivery limitations. To overcome this, we devised an internalized pump-catheter system for direct chronic convection-enhanced delivery (CED) into peritumoral brain tissue. Topotecan (TPT) by chronic CED in 5 patients with refractory glioblastoma selectively eliminated tumor cells without toxicity to normal brain. Large, stable drug distribution volumes were non-invasively monitored with MRI of co-infused gadolinium. Analysis of multiple radiographically localized biopsies taken before and after treatment showed a decreased proliferative tumor signature resulting in a shift to a slow-cycling mesenchymal/astrocytic-like population. Tumor microenvironment analysis showed an inflammatory response and preservation of neurons. This novel drug delivery strategy and innovative clinical trial paradigm overcomes current limitations in delivery and treatment response assessment as shown here for glioblastoma and is potentially applicable for other anti-glioma agents as well as other CNS diseases.

---

Glioblastoma, the most common primary brain malignancy, is uniformly fatal despite conventional therapy with surgery, radiation, and chemotherapy, underscoring the need for more effective treatments^1^. Promising anti-glioma drugs have failed clinically because of limitations in drug delivery and assessment of treatment response ^2,3^. We developed a chronic local drug delivery technique for gliomas that circumvents blood brain barrier limitations and avoids systemic toxicity to achieve effective intratumoral drug levels. We applied this technique in a clinical trial for recurrent glioblastoma patients using topotecan (TPT), a topoisomerase inhibitor that effectively kills proliferating glioma cells but is clinically impractical as a systemically delivered chemotherapeutic because of systemic toxicity and poor brain penetrance^4^. Our novel trial design enabled us to assess treatment effects using innovative cellular and genomic tissue analysis methodology that overcomes the limitations of conventional clinical metrics including radiographic response and survival as primary measures of efficacy.

Because glioblastomas are locally invasive, rarely metastasize, and usually recur within two centimeters of the original resection margin^5^, local drug delivery strategies have the potential to impact patient survival and provide insight into the direct effects of chemotherapy on the tumor microenvironment^6^. Convection-enhanced delivery (CED) is a method of local-regional drug infusion that delivers high concentrations of therapeutic compounds directly into the brain through a surgically implanted thin cannula attached to a microinfusion pump. Drugs are infused at flow rates that generate a positive hydrostatic pressure to distribute the infusate by bulk flow through the interstitial space. Because it circumvents blood-brain barrier limitations and avoids systemic toxicity, CED provides a pharmacokinetic advantage that is several orders of magnitude greater for maximizing drug levels in a targeted region of the brain compared to conventional diffusion-driven systemic methods such as oral or intravenous delivery^6,8-10^ .

We previously demonstrated the safety and feasibility of short-term CED in a clinical trial with topotecan for patients with refractory malignant gliomas^7^. However, a major shortcoming of previous CED approaches has been the difficulty in balancing the need to administer chronic treatment with the need to limit the treatment duration because of infection risks associated with externalized catheters and bedside pumps. To address this, we engineered a subcutaneously implanted catheter-pump system similar to one effectively used in Parkinson patients^11^. After successful preclinical modelling in a large animal^12^, we devised a clinical trial for patients with recurrent, refractory glioblastoma using chronic CED incorporating a refillable pump subcutaneously implanted in the abdomen for chronic and repeated intracerebral infusions of high-dose TPT chemotherapy. Sustained chronic delivery with multiple treatment cycles is important therapeutically as topoisomerase poisons such as TPT, like most chemotherapies, are cytotoxic to cycling cells in the S-phase where only a small percentage of glioma cells reside at any given time^13,14^. Preclinical studies in a rat glioma model with locally delivered TPT demonstrated improved survival when the infusion duration was extended to allow more tumor cells to cycle through the vulnerable S-phase^15^.

The goals of this clinical trial were to test the clinical utility of chronic CED in glioma patients, to assess the feasibility and safety of a subcutaneous pump/catheter construct, and to validate a non-invasive, radiographic method for monitoring drug distribution. Additionally, by collecting radiographically-localized biopsies of tumor and peritumoral tissue both before and after treatment, the trial design allowed us to overcome the limitations of radiographic and clinical response assessments conventionally used to determine chemotherapy effects in glioma trials^16,17^. This tissue collection provided an unprecedented opportunity to analyze both harmful and beneficial effects of chemotherapy on the tumor microenvironment utilizing sophisticated methodologies including: 1) advanced non-invasive radiographic imaging to monitor drug distribution and pharmacological effects; 2) extensive clinical monitoring for toxicity and quality-of-life metrics; and 3) comprehensive molecular, genomic, and cellular analyses of pre- and post-treatment tissue from the tumor and peritumoral regions. In addition to validating an improved drug delivery strategy, this trial design offers an innovative paradigm for clinical testing of new treatments for glioblastoma that overcomes current limitations in delivery and treatment response assessment.

## RESULTS

### Study Design and Patient Selection

The overall trial design consisted of multiple infusions of topotecan through a surgically placed intracerebral catheter connected to a subcutaneously implanted pump followed by radical resection of the tumor and removal of the pump/catheter four weeks later (**Figure 1A-B)**. Five patients with previously histologically confirmed malignant gliomas were enrolled in the trial, all of whom showed radiographic progression after surgical resection, temozolomide chemotherapy and external beam radiation. A preoperative MRI was performed to optimize selection of localized tumor biopsies and catheter trajectory to ensure optimal CED and tumor coverage. Patients underwent multiple stereotactic biopsies of the tumor and infiltrated brain tissue for comprehensive immunohistopathologic and molecular analyses as well as for comparison to post treatment specimens. One of the intratumoral biopsies was used for intraoperative histopathologic assessment to verify the diagnosis of recurrent tumor. A thin (1.5 mm outer diameter) silastic catheter was stereotactically positioned between the contrast enhancing tumor and the margin of the planned surgical resection to be performed at the end of the 4 week treatment (**Figure 1B**). This catheter placement strategy was designed to maximize drug delivery into the surrounding brain tissue and extend beyond the resection margin. The catheter was connected to silastic tubing that was tunneled subcutaneously then connected to a microinfusion pump implanted into the abdomen. Four treatment pulses were given consisting of 146 μM of TPT infused over 48 hours at 200 uL/min followed by a 5- to 7-day treatment holiday. Gadolinium was co-infused as a surrogate tracer during the first and fourth pulse and advanced non-invasive radiographic imaging was used to monitor drug distribution and pharmacological effects (**Figure 1C)**. After the fourth treatment pulse, the patient underwent surgical tumor resection along with removal of the pump and catheter system. Immediately prior to tumor resection, multiple radiographically-localized stereotactic-guided biopsies of tumor, tumor margin and surrounding invaded brain tissue were obtained providing a unique opportunity to analyze tissue for drug levels and treatment effects (**Figure 1D)**.

**Figure 1:**
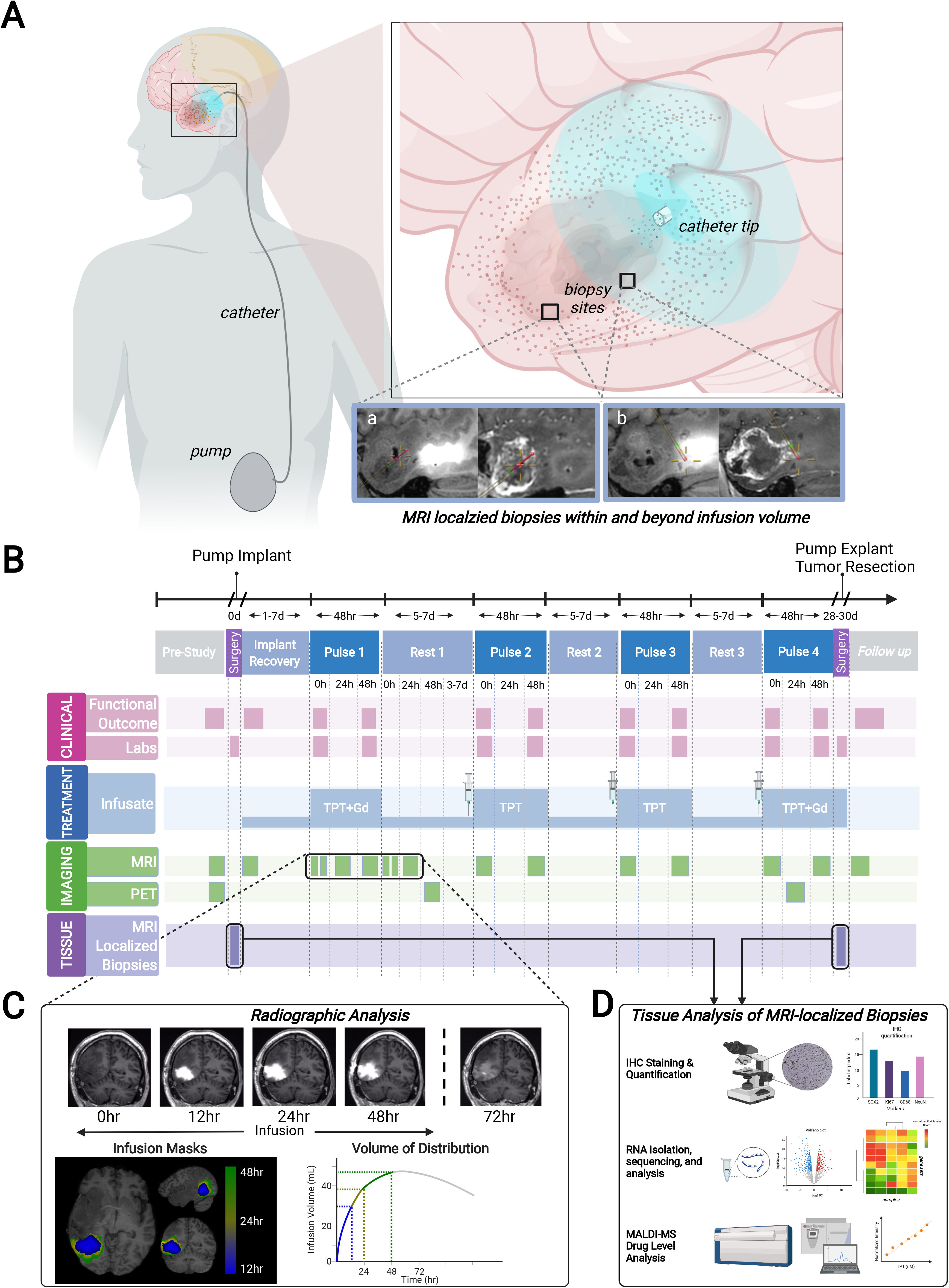
Study Overview and Timeline. **A:** Overview of the implantable chronic CED pump-catheter system, as well as drug infusion and acquisition of MRI-localized biopsies. A thin catheter was stereotactically placed midway between the tumor margin and the margin of the expected resection cavity, connected to silastic tubing that was tunneled subcutaneously and connected to a microinfusion pump implanted into the abdomen. MRI-localized biopsies were taken both at time of catheter implant and after treatment at explant. At explant, biopsies were taken both within and outside the volume of infused contrast. **B:** Schematic timeline describing the overall flow of the clinical trial. Patients enrolled in the trial underwent a baseline MRI and PET scan prior to undergoing stereotactically MRI-guided biopsies and implantation of the catheter with subcutaneous pump placement. TPT was delivered over 48 hours at 200 uL/mL in 4 cycles, with the pump refilled subcutaneously between each cycle, and 5-7 days of rest between infusions. MRIs were performed at set time points throughout the four cycles. Functional outcome measurements, physical exams, and basic lab panels were collected and assessed throughout the duration of treatment. At the completion of the 4 cycles the pump was explanted, further MRI-localized biopsies were acquired, and the tumor was resected. **C:** Several MRIs were performed during the first pulse of TPT+Gd to characterize volume of distribution of the infusate, as well as to build radiographically-defined maximum infusion masks for further analyses. **D:** MRI localized biopsies collected before and after CED underwent tissue analysis, including immunohistochemical staining and quantification, as well as RNA isolation, sequencing and analysis. On a small subset of these biopsies, MALDI-MSI was performed to quantify concentration of TPT in the tissue after infusion.

### Patient characteristics, tumor pathology and clinical outcomes

Basic demographic and clinical data of patients enrolled in the trial are summarized in **Table 1 and Supplementary Table S1**. One patient was excluded from the study when the diagnostic biopsy revealed histopathology consistent with pseudoprogression treatment effect without diagnostic evidence of recurrent tumor. Three male and two female patients (median age = 56; age range: 31 – 65 years) were enrolled in the trial at a median of 11 ± 14.8 months (range 6-42 months) from the time of initial diagnosis. All patients had biopsy-proven IDH1 wild type high grade glioma including one patient whose initial diagnosis was grade III astrocytoma. All other patients had initial diagnosis of glioblastoma. Two patients had a methylated MGMT promoter and one patient had an EGFRvIII mutation on initial diagnosis. All patients had previous treatments, including standard of care temozolomide chemotherapy and external beam radiation.^a^

**Table 1:** Basic clinical and pathological data from the five patients who completed the study. Further clinical information (including **Supplementary Table S1**) can be provided upon request to corresponding author.

| Patient | Sex | Age Range at enrollment | Initial Diagnosis | Tumor Location | Tumor Volume | IDH1 status | MGMT promoter methylation | Survival from Enrollment (months) |
| --- | --- | --- | --- | --- | --- | --- | --- | --- |
| Patient 1 | M | 31-35 | Anaplastic astrocytoma Grade 3 | R, Frontal | 1.9 mL | wt | Unmethylated | 12 |
| Patient 2 | F | 56-60 | GBM Grade 4 | R, Temporal | 12.9 mL | wt | Unmethylated | 17 |
| Patient 3 | M | 61-65 | GBM Grade 4 | L, Frontal | 5.1 mL | wt | Methylated | 10 |
| Patient 4 | M | 56-60 | GBM Grade 4 | R, Temporal | 18.0 mL | wt | Unmethylated | 5 |
| Patient 5 | F | 51-55 | GBM Grade 4 | R, Frontal | 4.1 mL | wt | Methylated | 25 |

Median survival for all 5 patients was 12 ± 7.6 months from time of enrollment (range: 5-25 months) and median overall survival from time of initial diagnosis was 23 ± 21.6 months (range: 13-68 months). Radiographic changes related to CED and surgical resection precluded a reliable determination of time to tumor progression based on RANO criteria^16^ **(Supplementary Figure S1A-C)**.

Overall, TPT by chronic CED was generally well tolerated and complications were uncommon and transient. Importantly, no significant systemic complications occurred **(Supplementary Figure S2)**. The most common treatment complaints were pain at the incision site (100%), fatigue (60%) and headache (40%), all symptoms grade 1 or 2 **(Supplementary Table S1)**. One patient had worsening of a baseline supplementary motor area syndrome during the initial infusion pulse. The syndrome improved over the ensuing treatment holiday and the remaining infusions were given at a 50% reduced infusion rate without further incident. Because this same patient also had a seizure between the 2^nd^ and 3^rd^ infusion pulse, the 3^rd^ infusion was reduced to 24 hours as a precaution given the patient’s pre-treatment baseline seizure disorder. This patient also developed transient SIADH which resolved with fluid restriction. Furthermore, this patient developed a lower extremity deep venous thrombosis during the treatment period, which was attributed to glioma associated coagulopathy^18^. An IVC filter was placed with no further complications.

All subjects were ambulatory and maintained their baseline Karnofsky Performance Score (KPS) throughout the TPT treatment **(Supplementary Figure S1D)**. The same patient described above had a transient decrease in KPS to as low as 70% due to a transient supplementary motor area syndrome, however KPS returned to baseline 90% at the end of Pulse 4. Quality of life (QoL) testing verified the safety and tolerability of the treatment as no significant changes were seen in the FACT-Brain QoL assessment, the FACIT Fatigue assessment or the PROMIS Global Health Measure **(Supplementary Figure S1F-G**). SAEs in the follow-up period associated with surgical resection or underlying disease but unrelated to the TPT treatment are reported in the Supplementary information.

### Chronic CED achieves large and stable volumes of distribution and delivers sustained therapeutic levels of TPT into and around the tumor

Using the MRI signal of co-infused gadolinium as a surrogate, **c**hronic CED resulted in a large and stable volume of drug distribution for all patients **(Figure 2A)**. Each patient’s time series was fit to a gamma function and the fits were averaged to estimate the maximal volume of distribution (mean = 20.4 ± 10.5 mL; range = 12.0 - 35.9 mL) and time to peak volume after start of infusion (mean = 43.1 ± 11.5 hrs, range = 26.8 - 55.5 hrs) **(Figure 2B)**. Backflow was a small fraction (8.8%) of total infused volume (maximum backflow volume: mean = 1.8 ± 2.1 mL; range = 0 - 4.5 mL) **(Supplementary Figure S3)**. Mass spectrometric imaging (MALDI-MSI) was performed to quantify TPT concentration on 12 MRI-localized biopsies taken post-CED to determine the presence of therapeutic drug levels within the peritumoral volume. Eleven of 12 biopsies had detectable levels of TPT with pixel values above the LOD (3.2 uM) (**Supplemental Figure S4A)** and average TPT concentrations ranging from 1.1μM to 30μM (mean = 8.9 ± 2.3 μM). Micromolar concentrations of TPT were present up to 6 centimeters from the catheter tip and in all biopsies taken within the maximum volume of gadolinium distribution (**Supplemental Figure S4B**).

**Figure 2:**
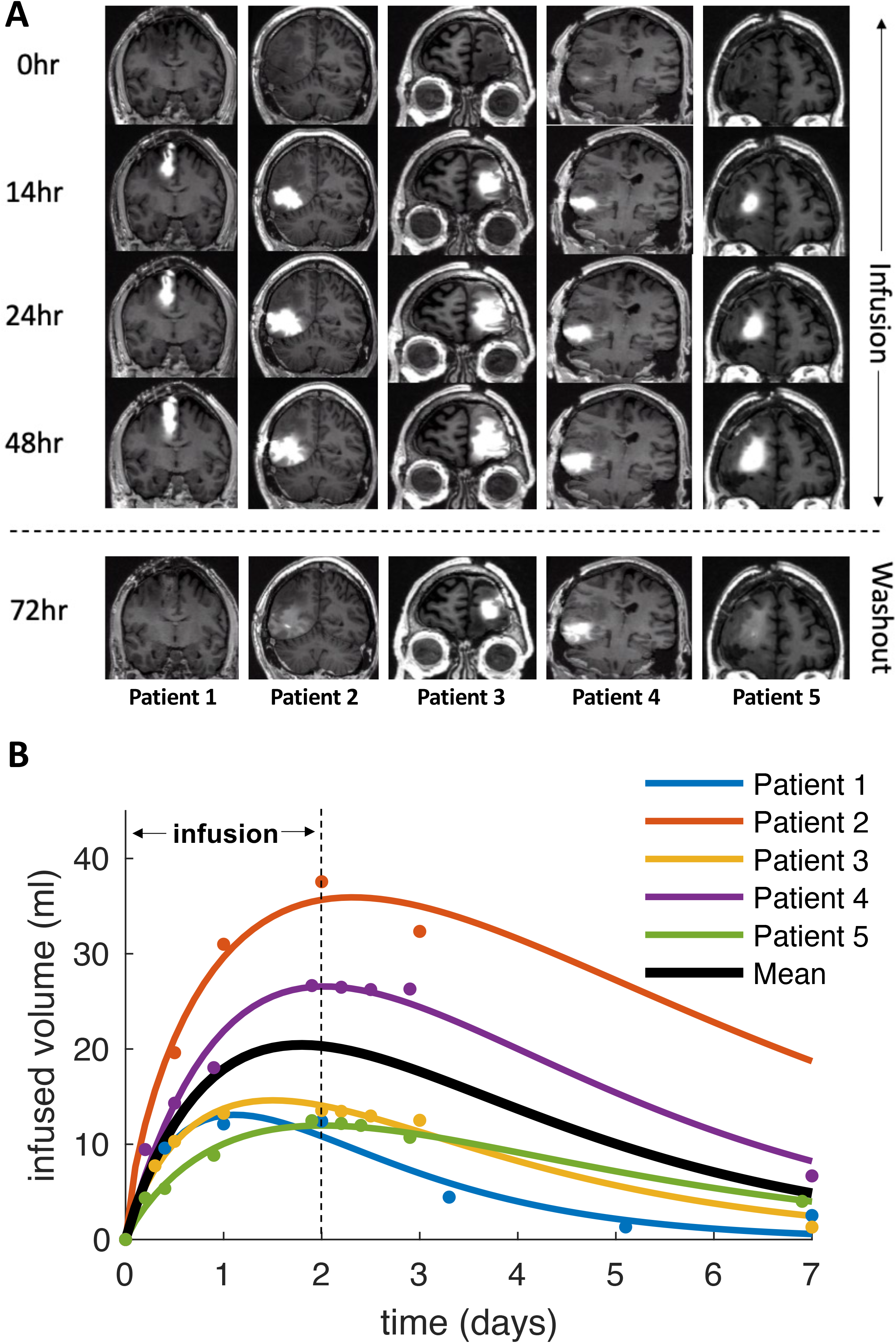
Chronic CED of Topotecan achieves large and stable volumes of distribution. **A:** Each patient was infused over the course of 48 hours, after which the washout period began. All five patients showed large changes in infused contrast. **B:** The volume of the infused contrast was plotted as a function of time and fit to a gamma function for each patient. The solid black line represents the mean time course across subjects, which peaked at 43.1 hours with a mean volume of 20.4 mL.

### Chronic CED of TPT effectively targets proliferating tumor cells, and shifts glioma phenotype

Tissue collected from patients pre- and post-treatment was analyzed by immunohistochemistry to determine the effects of TPT on tumor cells and surrounding microenvironment **(Figure 3A-D)**. Tumor burden was assessed by staining for SOX2, a highly pervasive glioma cell marker that is expressed by the vast majority of tumor cells in GBM^19^. The SOX2 labeling index was significantly decreased in post-CED biopsies compared to pre-CED biopsies (18.5% vs. 25.8%; p=0.03). Additionally, Ki67 proliferation index was significantly lower in post-CED biopsies compared to pre-CED biopsies (1.4% vs 3.9%; p=0.009).

**Figure 3:**
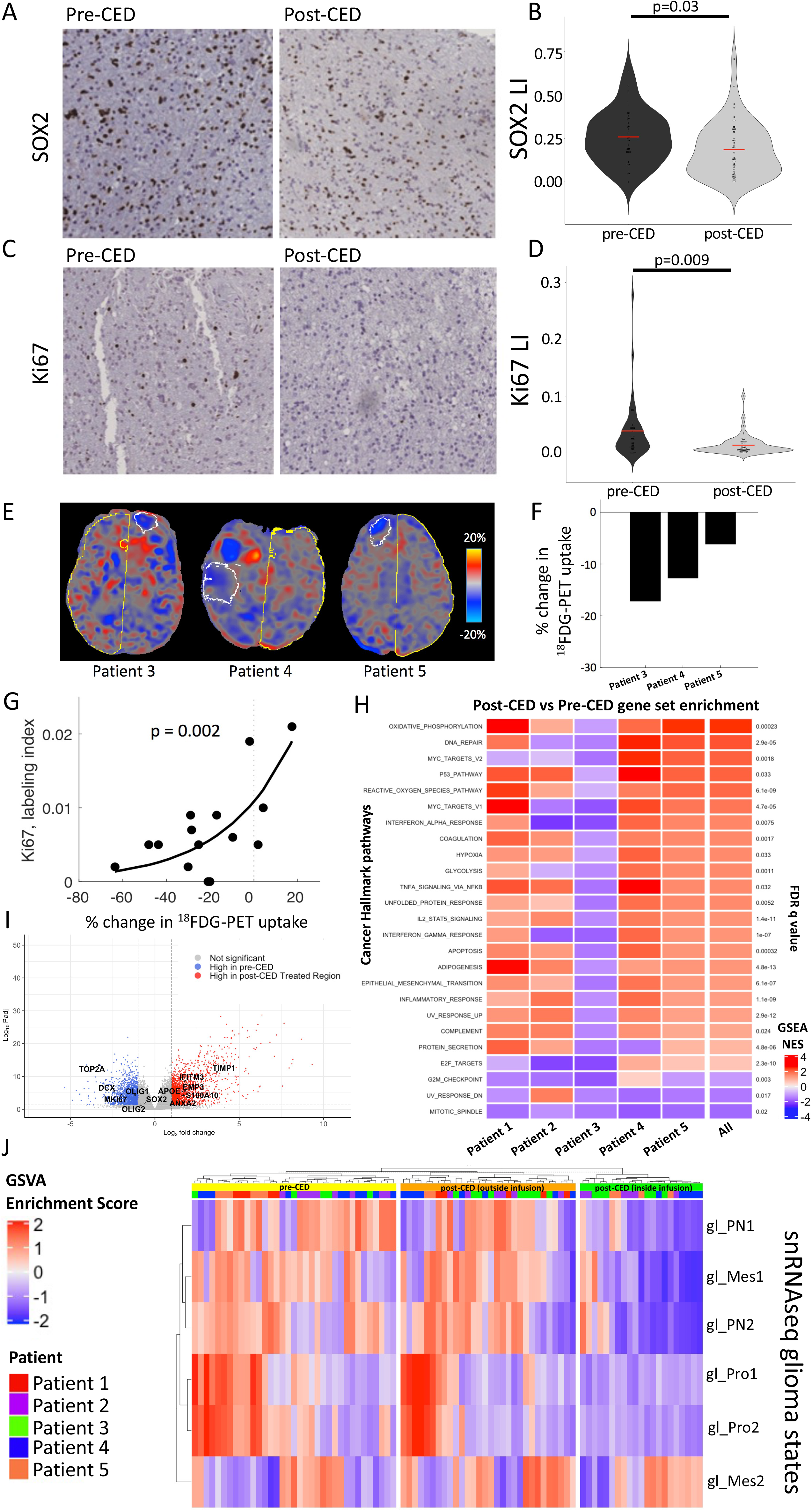
Chronic CED of Topotecan targets proliferating tumor populations and shifts tumor phenotype. **A-D**: Representative images pre-CED and post-CED immunohistochemically stained for SOX2 (panglioma cell marker) and Ki67 (proliferation marker), respectively, Both stains have corresponding violin plots, which display quantification of each marker by labeling index across all MRI-localized biopsies from all patients (n=87), comparing biopsies taken pre- and post-CED using a student’s T-test. **E:** The difference between post- and pre-infusion PET images were computed and converted to percent signal change. The white outline represents the maximum infused volume; the yellow outline represents the control hemisphere. Blue voxels represent decreases in metabolism, red voxels represent increases in metabolism, and gray voxels represent no change after treatment. **F:** All three patients showed a large (6.2-17.2%) reduction in metabolism within the infused volume mask. **G:** The regions treated with TPT demonstrated reduced Ki67 labeling and reduced ^18^FDG metabolism. These two measures were exponentially related, such that, the larger the reduction in metabolism, the lower the proliferation index. **H**: Heatmap displaying patient-by-patient gene set enrichment analysis (GSEA) comparing MRI-localized biopsies before and after treatment for MSigDb “Cancer Hallmarks” ontologies. Red indicates a pathway is enriched post-CED, while blue indicates the pathway is enriched pre-CED. Significantly enriched gene sets across all patients via Fisher integration are displayed on the left of the heatmap, with the corresponding adjusted FDR q-value to the right. **I:** Volcano plot displaying differentially expressed genes between pre-CED and post-CED MRI-localized biopsies within volume of treatment across all 5 patients (n=56). Cutoffs were log-2-fold-change > |1| and adjusted p-value<0.05. Select significantly differentially expressed lineage-associated genes are marked. **J**: Heatmap displaying GSVA analysis of MRI-localized biopsies for six snRNAseq-derived glioma cell state signatures. GSVA scores for a given gene set are scaled across all samples. On the top of the heatmap, each biopsy is annotated by patient, as well as whether the biopsy was taken pre-CED or post-CED. The post-CED biopsies were further radiographically stratified by whether they were inside or outside the maximum volume of distribution.

To further assess the functional impact of chronic infusion of TPT, 3 of the 5 patients underwent ^18^FDG-PET imaging before and after CED treatment. The maximal volume of the infused gadolinium was used to calculate the percent signal change in ^18^F-FDG metabolism. ^18^FDG-PET is a surrogate of glucose metabolic activity and linked to proliferation status (**Figure 3E, white outline)**. All three patients showed a significant reduction (Patient 3 = -17.2%; Patient 4 = -12.7%; Patient 5 = -6.2%) in glucose uptake within the treated regions (**Figure 3F)**. Proliferating glioma cells have been shown to heavily rely on glycolysis ^20^. To further assess the relationship between the treatment-induced alterations in metabolism with proliferation, we analyzed correlations between KI67 proliferation index and change in FDG uptake after treatment across MRI-localized biopsies taken post-CED. Biopsies that showed a treatment-related decrease in glucose uptake also showed a reduction in the Ki67 labeling index. Across all biopsies, a significant exponential relationship was seen between the magnitude of treatment-related reduction in metabolism and proliferation (p = 0.002) (**Figure 3G)**. Therefore, FDG-PET studies can be used in GBM patients undergoing chronic CED of chemotherapy to track effects on the proliferating tumor cell population in real time.

To further understand the effects of TPT on tumor proliferation, RNA sequencing and differential gene expression analysis were performed on select MRI-localized biopsies taken pre-CED at time of catheter placement (n=35) and post-treatment at the time of tumor resection (n=51) (**Supplementary Table S2)**. Gene set enrichment analysis (GSEA) demonstrated marked heterogeneity across patients pre- and post-treatment but demonstrated clear patterns of tissue response, with increases in gene signatures for DNA damage, apoptosis, and generation of reactive oxygen species, as well as upregulation of several metabolic programs, including oxidative phosphorylation (**Figure 3H, Supplementary Table S3)**. Mitotic spindle and other proliferation-associated gene ontologies were significantly decreased across all patients, further demonstrating the impact of chronic TPT infusion on the proliferating tumor cell population. Of note, unlike the other four patients in the study, Patient 3 demonstrated marked downregulation of all depicted ontologies post-treatment.

To understand the effects of TPT infusion on specific subpopulations of glioma cells, single-sample gene set enrichment analysis (GSVA) was performed on all 86 MRI-localized biopsies using gene signatures derived from snRNAseq for distinct glioma cell states ^21^, including proliferative states (gl_Pro1 and gl_Pro2), Proneural states (gl_PN1 and gl_PN2) and Mesenchymal states (gl_Mes1 and gl_Mes2) (**Figure 3J)**. In order to fully ascertain the spectrum of treatment effects, post-CED biopsies were further stratified by whether they were inside or outside the maximum volume of distribution, as described above. When comparing post-CED biopsies taken from within the treatment volume to the pre-CED biopsies, there was a significant shift in the enrichment scores for these glioma cell states, with a significant decrease in the samples showing highest enrichment for the proliferative or proneural signatures and a significant increase in the samples showing highest enrichment for mesenchymal signatures (p<0.001). Notably, the majority of post-CED samples from within the treatment volume showed the highest enrichment for the gl_Mes2 snRNAseq signature, and none showed highest enrichment for the proliferative signatures (gl_Pro1 and gl_Pro2). This analysis was repeated using glioma cell state gene sets derived by single-cell RNA sequencing (scRNAseq) as reported by Neftel et al^22^, also demonstrating a significant enrichment (p=0.01) of mesenchymal signature after treatment (**Supplementary Figure 5A)**, Canonical mesenchymal genes were significantly upregulated post-CED *within* the treatment volume, and proliferation-associated and glioma progenitor genes were significantly negatively differentially expressed (**Figure 3I, Supplemental Table S5)**. In contrast, comparing the pre-CED samples to the post-CED samples taken outside the treatment volume did not show a significant change in the distribution of enrichment scores for the glioma cell state signatures(p=0.07). These results show that chronic CED of TPT causes a significant depletion of proliferating/proneural signatures and an increase in mesenchymal signatures selectively within the treatment volume.

### Chronic CED of TPT affects the tumor microenvironment

Given the shift toward a mesenchymal signature seen in the post-CED samples, and the previously described association between mesenchymal GBM and inflammatory cells^19,21,23^ we analyzed the effects of chronic CED of TPT on the tumor microenvironment. CD68, a macrophage marker and surrogate for inflammation was significantly increased in post-CED biopsies as assessed by immunohistochemistry (17.4% vs. 9.6%; p=0.02) (**Supplemental Figure S5B-C**). Post-CED biopsies were also positively enriched in pro-inflammatory transcriptional programs, including interferon alpha response, interferon gamma response, IL2-STAT5 signaling and TNF-NFKB signaling (**Figure 3H**). Differential gene expression analysis was performed by pooling biopsies across all patients, and pre-CED versus post-CED comparison demonstrated significant increase in several pro-inflammatory cytokines and other immuno-active markers **(Supplemental Figure S5D)**, including CCL18, CCL20, CCL22 and CXCR1. Overall, this demonstrates that chronic TPT infusion leads to global changes affecting both the tumor cell phenotype and the tumor microenvironment. Lastly, to assess the effects of chronic TPT infusion on the neuronal population we performed immunostaining with the neuronal marker NeuN (not shown). NeuN labeling index demonstrated variability across samples, depending on biopsy location, but showed no significant difference in post-treatment biopsies as compared to pre-treatment biopsies, providing supportive evidence that TPT is not significantly toxic to neurons (post-CED 5.4% vs. pre-CED 4.7%; p=0.770) (**Supplemental Figure S5E-F**).

## DISCUSSION

Our successful use of a subcutaneously implanted pump to provide prolonged drug infusion duration overcomes a significant shortcoming in the clinical efficacy of CED. We used this system to treat refractory glioblastoma patients, delivering high doses of TPT directly into the tumor and surrounding brain over 4 weeks without serious neurological or neurobehavioral events thereby circumventing the limitations associated with conventional systemic delivery. Using MRI to non-invasively monitor the distribution of co-infused gadolinium as a surrogate for TPT distribution, we demonstrated large and stable volumes of drug distribution.

The advantage of an implantable CED pump allows for percutaneous refilling and the potential for sequential or simultaneous treatment algorithms using one or more drugs while maximizing distribution volume. The pump/tubing/catheter construct that we utilized is improvised from a variety of different commercial sources and is designed to be used with a skill set common to all neurosurgeons. The tested system has numerous features designed to ease its clinical use: 1) self-contained subcutaneously implanted hardware to avoid infection and facilitate chronic use; 2) implanted microinfusion pump with a reservoir containing the treatment drug which can be percutaneously refilled or emptied with a needle and syringe; 3) wireless programming pump technology to modify flow rate; 4) simple flexible thin bore catheter which can be implanted precisely with stereotactic guidance and can accommodate flow rates within the therapeutic range without significant backflow; 5) incorporation of a technique for co-infusing gadolinium to monitor volume of delivery in real time using a safe and reliable non-invasive methodology; and 6) safety profile suitable for use in an out-patient setting. Improvements in the CED construct are a source of ongoing study by our lab and others including catheter design, use of multiple catheters, as well as improvements in pump technology, monitoring software and stereotactic technologies.

While validating the safety and feasibility of chronic CED, our unique treatment protocol provides a novel broad clinical framework to study the effects of locally delivered therapy. Performing a series of pre- and post-therapy MRIs and PET scans, as well as taking dozens of MRI-localized biopsies upon implantation and removal of the CED pump-catheter system, facilitated the study of clinical intervention from a lens typically reserved for preclinical animal models. In this study, we used imaging to monitor the volume of distribution of the infused therapeutic, as well as to examine the effects of treatment on the tumor and its microenvironment. Image-localized biopsies before and after treatment allowed us to study the effects of TPT on the tumor and its surrounding environment via a multi-omics approach. This is a useful and strategic approach to study the tissue-specific effects of novel therapies to the brain in a direct clinical setting with ever greater utility, not only for gliomas but other non-neoplastic CNS diseases as well^11,24,25^, especially as next-generation methods of analyzing tissue evolve, such as single cell transcriptomics, proteomics, metabolomics and lipidomics.

Comparison of pre-and post-treatment tissue demonstrated consistent responses to topotecan after treatment. Post-treatment tissue saw a greater than 2.5-fold reduction in Ki67 labeling index and decreased tumor burden evidenced by a 1.5-fold reduction in SOX2 labeling index, with a relative preservation of neurons. The decreased proliferative signature was further verified with bulk RNA sequencing analysis, demonstrated by negative enrichment of curated proliferative gene ontologies. Single sample GSVA analysis using snRNAseq-derived glioma cell state signatures demonstrated marked reduction of the proliferative signatures, and a corresponding increase in mesenchymal and inflammatory signatures. Notably, while previous studies have shown that mesenchymal signatures are frequently enriched in post-treatment recurrent GBM^26,27^ our analysis revealed an mesenchymal shift in samples taken immediately after treatment, presumably months prior to tumor recurrence, supporting the idea that the mesenchymmal signature represents the cells that escape treatment and will eventually give rise to recurrence. We also detected a marked inflammatory response in the tumor microenvironment. CD68 labeling index was increased after treatment, indicating increased proportions of activated myeloid cells, including microglia and macrophages. Furthermore, GSEA demonstrated upregulation of inflammatory transcriptional programs after treatment, indicating the likelihood of increased immune infiltrate into the tumor bed and activation of the local innate immune response. While the clinical significance of this robust inflammatory response remains unclear, our analysis identified increase in several cytokines that have been associated with glioma growth and poor outcome^28-30^. Future studies could test if targeting this proinflammatory, mesenchymal glioma subpopulation, perhaps in combination with topotecan or another S-phase poison, could enhance responses to therapy and even prolong survival.

Overall, this clinical study provides a novel experimental framework from which to design future studies, not only for gliomas, but other CNS diseases as well. First, the feasibility of the internalized pump-catheter system for CED has important implications for both expanding the duration (potentially indefinitely) and repertoire (multi-drug/sequential drug regimens) of therapy, supported by the ability to non-invasively monitor drug delivery. Additionally, because the blood brain barrier is bypassed, new classes of drugs and targeted compounds can be exploited including high molecular weight compounds, proteins, viruses, liposomes, nanoparticles and other biologics that would not be feasible with systemic delivery due to toxicity or metabolic breakdown. Second, MRI localized biopsies taken before treatment at pump implantation can potentially allow tailoring of the treatment regimen to each patient’s tumor, and even implement combination therapies to target distinct glioma subpopulations. Lastly, biopsies taken immediately after therapy provide an unparalleled view into mechanisms of tumor resistance and recurrence from which new therapeutic approaches can be designed.

## ONLINE METHODS

### Patients

Eligible patients were ≥18 years of age with previously histologically confirmed malignant glioma (WHO grade III-IV) treated with surgical resection, temozolomide chemotherapy, and external beam radiation showing clinical evidence of recurrent glioblastoma based on increasing contrast enhancement on MR or CT imaging while on stable or increasing dose of steroids. Two patients received prior immunotherapy, Pembrolizumab and Nivolumab respectively. One patient received the tyrosine kinase inhibitor Neratinib as part of a clinical trial. Additional eligibility criteria included Karnofsky performance status ≥ 70 and stereotactically-accessible supratentorial contrast-enhancing tumor localized to a region < 32 cc in volume on pre-enrollment MRI.

All patients gave written informed consent to the protocol which was approved by the Columbia University Irving Medical Center Institutional Review Board and the Food and Drug Administration. The Principal Investigator (JNB) was the sponsor of the Investigational New Drug approval from the Food and Drug Administration application (IND 131889) for topotecan and the catheter/pump delivery system.

### Study Design

Treated patients underwent multiple stereotactic biopsies of the tumor immediately before treatment to verify the diagnosis and distinguish active tumor from radiation necrosis or pseudoprogression. Biopsies were taken from within the tumor, the tumor margin and surrounding invaded brain tissue. One patient was removed from the study when biopsy failed to confirm active tumor. A 1.5 mm outer diameter silastic Spetzler lumbar shunt catheter (Integra, Plainsboro, NJ) was stereotactically placed adjacent to the tumor to optimize the drug infusion into the peritumoral invaded brain tissue based on pre-operative MRI planning. The catheter was connected to silastic tubing that was subcutaneously tunneled and connected to a SynchroMed II infusion pump (Medtronic, Minneapolis, MN) implanted in the abdomen. The infusate consisted of 1:100 gadolinium (Gadavist, GE Healthcare, Marlborough, MA) plus 146 uM TPT (Hycamtin, GlaxoSmithKline, research Triangle Park, NC).

Active infusion was initiated 1-7 days after pump and catheter implant to allow time for expected and routine recovery from surgery. The patients underwent four 48-hour pulses of therapy interspersed by 5-7 day therapy holidays. Notably, gadolinium was added to the TPT infusate only during the first and the fourth CED pulses. Rationale for 48-hour pulsatile infusion was derived from preclinical porcine studies demonstrating that the largest relative gains in volume of distribution of infusate occur within the first 24-48 hours before declining as infusions reach a steady state^12^. Similarly, the total volume of infusion and dose concentration was based on our previous clinical trial in which 40mL of drug was infused and a maximum tolerated dose of 218uM TPT was defined^7^. Pumps were emptied, then refilled percutaneously prior to each 48-hour pulse. In each pulse cycle, patients received 200uL/hr of TPT for 48 hours (daily volume = 4.8 mL; total volume = 9.6 mL) followed by between five to seven days of therapy holiday during which the pumps were set at the minimum programmable rate (2uL/hr; 5-7 day volume = 0.24-0.34mL). This cycle was repeated two more times. After three complete cycles, patients received a fourth and final 48-hour pulse. After the fourth dose, patients were taken to the operating room for catheter/pump removal and tumor resection.

### Evaluation of safety

Patients underwent daily neurological examinations during the treatment period and at continued periodic time points following treatment: 1-2 weeks, 4-6 weeks and then every 1-3 months. QoL and neurocognitive testing using Cognitive Stability Index or the Physical or Mental QoL assessments were immediately performed before and after each treatment pulse.

### Tissue collection

Tumor and peritumoral brain tissue samples were collected at two time points: 1) at surgery for implantation of pump and catheter prior to drug infusion; and 2) following completion of fourth and last TPT infusion, immediately prior to tumor resection and pump/catheter removal. Stereotactic trajectory planning using the iPlanet BrainLab (Version 3.05) was used to maximize the number of biopsies from the fewest trajectories. An average of 7 biopsies were taken during catheter/pump implantation; an average of 11 biopsies were taken during pump removal and surgical resection. Biopsies were taken from different areas within the tumor and peritumoral tissue (in areas of non-eloquent brain tissue) at variable distances from the catheter tip location. The stereotactic nature of biopsies allowed for classification of biopsies to contrast enhancing/non-enhancing and to within convection area or not. Additional gross tissue specimens were collected during surgical resection and these were roughly categorized based on anatomical features and location from catheter tip. All biopsies were divided into 2 pieces in the neuropathology reading room, adjacent to the operative suite. One piece was immediately flash frozen in liquid nitrogen for RNA sequencing and drug analysis. A second piece was fixed in 10% (v/v) formalin and subsequently paraffin embedded for histologic analysis.

### MR & PET Imaging acquisition

MR imaging was performed on a 3T GE Signa HDxt scanner (software version HD23.0_V03_1614.b). Sagittal volumetric gradient echo T1-weighted (T1w) MRIs (TR = 7.27 ms, TE = 2.7, inversion time = 450 ms, matrix = 224 × 512 × 512, voxel = 0.7 × 0.47 × 0.47 mm) were obtained pre-operatively and on post-operative day 0 or 1 following catheter implantation to confirm catheter placement prior to infusion initiation. For the first infusion, MRIs were collected ∼8 hours, ∼14 hours, ∼24 hours, and ∼48 hours after the start of infusion, and ∼8 hours, ∼14 hours, and ∼24 hours after completion. For the remaining infusions, MRIs were collected immediately prior to and ∼48 hours after the start of each infusion. ^18^F-FDG PET imaging was performed on a Siemens Biograph 64_mCT (software version VG60A; matrix = 400 × 400 × 148, voxel = 1.0 × 1.0 × 1.5 mm) pre-operatively, ∼48 hours after start of pulse 1, and ∼24 hours after start of pulse 4.

### MR & PET Imaging analysis

MRI data were processed with FMRIB Software Library (FSL version 6.0.0) ^31^ and Matlab (2020b, MathWorks) software. T1w and PET images from all scanning sessions were co-registered to the pre-implantation T1w scan using FSL-FLIRT (linear, 6 degrees of freedom). T1w images were intensity normalized using histogram matching to a healthy control subject. To calculate the spatial distribution of the infused gadolinium contrast, the pre-implantation T1w image was subtracted from all subsequent T1w images. The difference images were then thresholded and binarized to create a 3D mask of the infused volume at each time point and the mask volumes were then plotted as a function of time for each patient. To estimate the time to peak volume and maximum achieved volume, the time course for each patient’s first infusion was fit to a three-parameter gamma function

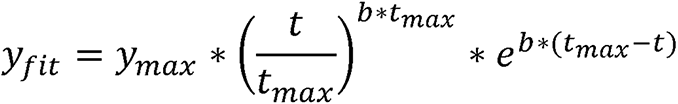

where *y*_*max*_ represents the peak volume, *t*_*max*_ represents the time of peak volume and *b* represents the shape of the function. To estimate backflow from the catheter, a mask was drawn around the infused volume located in the subdural CSF at the base of the catheter and the volume plotted as a function of time. All ^18^F-FDG-metabolism images were intensity normalized to the patient’s pre-infusion, contralesional hemisphere using histogram matching. Post-infusion ^18^F-FDG-metabolism images were converted to percent signal change by dividing by the pre-infusion image. The maximum infusion mask was used to estimate the effect of TPT on ^18^F-FDG metabolism.

### Measurement of TPT concentrations

TPT concentrations were measured in tissue samples using matrix-assisted laser desorption/ionization mass spectrometric imaging (MALDI MSI) methodology.

#### MALDI tissue preparation

A tissue microarray (TMA) with 1.5 mm core diameter channels composed of 40% gelatin was used to create a concentration ladder of TPT. To account for the tissue matrix, human autopsy brain homogenates were spiked with varying concentrations of TPT ranging from 1 - 50 μM. The spiked homogenates were punched into the TMA channels and frozen at -80 °C. Brain specimens and TMA were sectioned at 10 μm thickness, and thaw mounted directly onto indium tin oxide (ITO) slides. Serial sections were used for MALDI MRM MSI and brightfield microscopy imaging (Zeiss Observer Z.1, Oberkochen, Germany) of hematoxylin and eosin (H&E) staining. To avoid TPT interconversion between the lactone and carboxylate forms, a low pH phosphate solution was sprayed first on the tissues using a TM-sprayer (HTX imaging, Carrboro, NC). Two coats of the phosphate buffer solution (pH 3) was sprayed at a flow rate (0.05 mL/min), spray nozzle velocity (1200 mm/min), nitrogen gas pressure (10 psi), spray nozzle temperature (80 °C), and track spacing (2 mm) were sprayed on the tissue samples. The MALDI matrix 2,5-dihydroxybenzoic acid (160 mg/mL) was dissolved in 70:30 methanol: 0.1% TFA with 1 % DMSO. The ITO slides with tissue sections coated with phosphate solution were sprayed with the matrix using a TM sprayer with a two-pass cycle at a flow rate (0.18 mL/min), spray nozzle velocity (1200 mm/min), nitrogen gas pressure (10 psi), spray nozzle temperature (75 °C), and track spacing (2 mm).

#### MALDI mass spectrometry imaging

Matrix assisted laser desorption ionization (MALDI) MS imaging and quantitation of TPT from clinical specimens was performed using a timsTOF fleX mass spectrometer (Bruker Daltonics, Billerica, MA) operating in positive ion mode. A multiple reaction monitoring (MRM) method was applied to improve sensitivity and selectivity for quantitation. Using the ESI configuration on the dual-source instrument, a method for each drug was developed in which the ion transfer funnels, quadrupole, collision cell, and focus pre-TOF settings were optimized using TPT. Calibration of the mass range for each method was performed through the direct infusion of an Agilent tune mix solution (Agilent Technologies, Santa Clara, CA) and the MRM settings were optimized with direct infusions of TPT. The optimal collision energy of 20 eV and 50 eV for TPT, were determined, respectively. In both cases, the isolation width was set to 3 *m/z*. The mass range was selected to encompass both the precursor and product ions and was set to *m/z* 300-600 for TPT. For MALDI MSI quantitation, the transition of precursor to product was *m/z* 422.107377.111 for TPT. A spatial resolution of 100 μm was implemented for both MALDI MSI sequences. However, the repetition rate of the laser and laser shots per pixel varied for TPT (3,000 laser shots at 5,000 Hz). SCiLS Lab software (version 2020a premium, Bruker Daltonics, Billerica, MA) was used for data visualization, and total ion current (TIC) normalization was applied to the dataset. The limit of detection (LOD) and limit of quantification (LOQ) were based on signal-to-noise (S/N) ratio of > 3 and > 10 respectively. A linear regression of ion intensity and concentration were plotted to calibrate the MALDI signal intensity for quantitation of TPT.

### Immunohistochemistry

Paraffin embedded tissue was serially sectioned into 5μM sections mounted on immunoblank slides. Immunoperoxidase was performed using Antibodies to Sox2 (Rabbit, abcam #ab92494), Ki67 (Rabbit, Cell Signaling #9129), CD68 (Mouse, abcam #ab955), Neun (Mouse IGg1, Millipore #MAB377) under a standard staining protocol with deparaffinization, antigen retrieval using 10mM sodium citrate buffer, Vectastain ABC Kit (PK4001 for rabbit, PK4002 for mouse) and Dako DAB+ liquid substrate (#K3468). After peroxidase staining, slides were immersed in dH_2_O, counterstained with hematoxylin, dehydrated and coverslips were mounted using Permount mounting medium (Fisher Chemical #SP15-100).

### Immunohistochemistry quantification

Stained slides were scanned and digitized using a Leica SCN400 automated image digitizing system. A total of 87 MRI-localized biopsies were quantified for each stain, including pre-CED n=37) and post-CED (n=50). For each SCN file, ImageJ was used to outline tissue area at 20x magnification. Semi-automated quantification of DAB stained cells and nuclei was performed as follows. Images were split into blue channels for nuclei quantification and red channels for DAB quantification. Images were converted to 8-bit, normalized and thresholded using the local thresholding algorithm first described by Phansalkar et al.^15^ The local threshold is determined using the following algorithm: t=mean*(1+p*exp(-q*mean) + k*((stdev/r)-1)) where mean and STD are calculated from a 3×3 square of pixels centered on the pixel of interest. Values of k, r are optimized based off of multiple high-powered fields of masks ensuring accurate quantification on each image. Concordance between manual and semi-automated quantification was confirmed for each stain with r^2^ value greater than 0.90 in images from each patient before application to all biopsies. Labeling indices were calculated from dividing DAB+ cells over total number of cells (nuclei).

### RNA isolation, quantification and sequencing

Biopsies (n=86) were lysed using a 5mm stainless steel bead (QIAGEN, 69989). RNA was extracted from tissue lysate using the RNeasy Mini kit (QIAGEN, 74106). Only samples with RNA Integrity Number (RIN) greater than 6 were used for sequencing.

Expression profiles were generated using the Illumina TruSeq v2 RNA-Seq kit with 80 million paired end base reads on a Novaseq 6000 sequencer. Alignment and mapping of RNA sequencing reads was done in partnership with the Columbia Sulzberger Genome Center. Transcripts were pseudo-aligned, mapped and quantified to counts using the Kallisto pipeline.^16^

### RNAseq normalization, Differential gene expression analysis, and gene set enrichment analysis

All downstream RNA sequencing analysis was performed in the statistical programming language R (version 4.0.3). Raw transcript counts for each sample were normalized via the “DESeq2” package after filtering out genes that are not protein-coding. Differential gene expression analysis was also performed via DESeq2, comparing post-treatment biopsies (n=51) to pre-treatment biopsies (n=35), both on a patient-by patient level, as well as by pooling samples across all patients ^32^. Volcano plot and heatmap were generated based on normalized expression between pre-and post-CED biopsies, generated via “EnhancedVolcano” package^33^. Gene Set Enrichment Analysis (implemented via the GSEA desktop version 4.1.0) was performed using the 50 Cancer Hallmarks gene sets patient-by patient comparing pre-treatment and post-treatment MRI-localized biopsies. Fisher p-integration was performed on FDR-adjusted p-values across all patients for each pathway, and significant pathways (p<0.05) were visualized on a heatmap with normalized enrichment scores for each patient.

### Glioma State Analysis

GSVA analysis^34^ was performed on all MRI-localized biopsies (86 samples) using gene signatures for six distinct glioma cell states derived from single-nuclei RNA sequencing (snRNAseq) ^21^, as well as scRNAseq signatures derived from Neftel et al, and a heatmap was generated displaying the enrichment scores for each gene set for each biopsy, scaled across all samples. Each biopsy was annotated by the glioma cell state it was most enriched in (**Supplementary Table 4**), and Pearson’s chi-squared test was performed to statistically quantify change in phenotype between conditions.

## Data Availability

All data produced in the present study are available upon reasonable request to the authors

## Acknowledgements

We would like to acknowledge the Khatib Foundation, the Irving Institute for Clinical and Translational Research, the Herbert Irving Comprehensive Cancer Center Molecular and Pathology Core, and the patients, nurses, technicians and physicians who have made this work possible

## SUPPLEMENTARY INFORMATION

### Supplementary Figure Legends

**Supplementary Figure S1.**
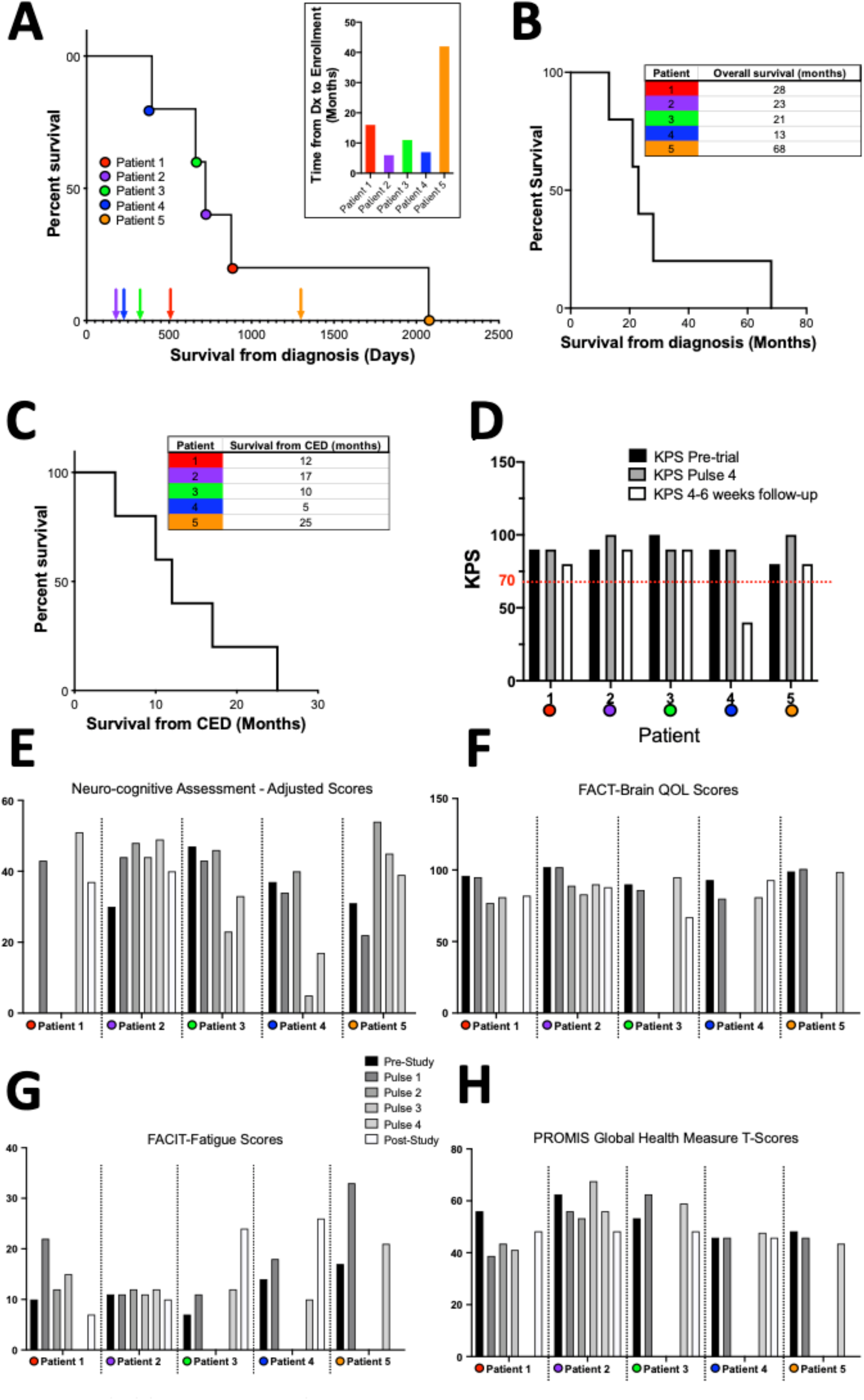
**A:** Kaplan Meier curve for all patients demonstrating overall clinical course for all patients. Arrows indicate time point of enrollment in the clinical trial while circles indicate time of death. Each patient is color coded. Inset: bar graph showing time from diagnosis to enrollment in the trial. **B-C:** Kaplan Meier curves and tables demonstrating survival from initial diagnosis/overall survival and survival from initiation of CED TPT. **D:** Bar graphs showing KPS scores pre-trial, at the end of Pulse 4 and at 4-6 weeks follow-up. Red line marks a KPS of 70% considered to be the cutoff for significant changes in functional status. **E:** Bar graphs showing neurocognitive assessments throughout treatment. **F-H:** Bar graphs showcasing QoL assessment scores throughout treatment for all 5 patients.

**Supplementary Figure S2.**
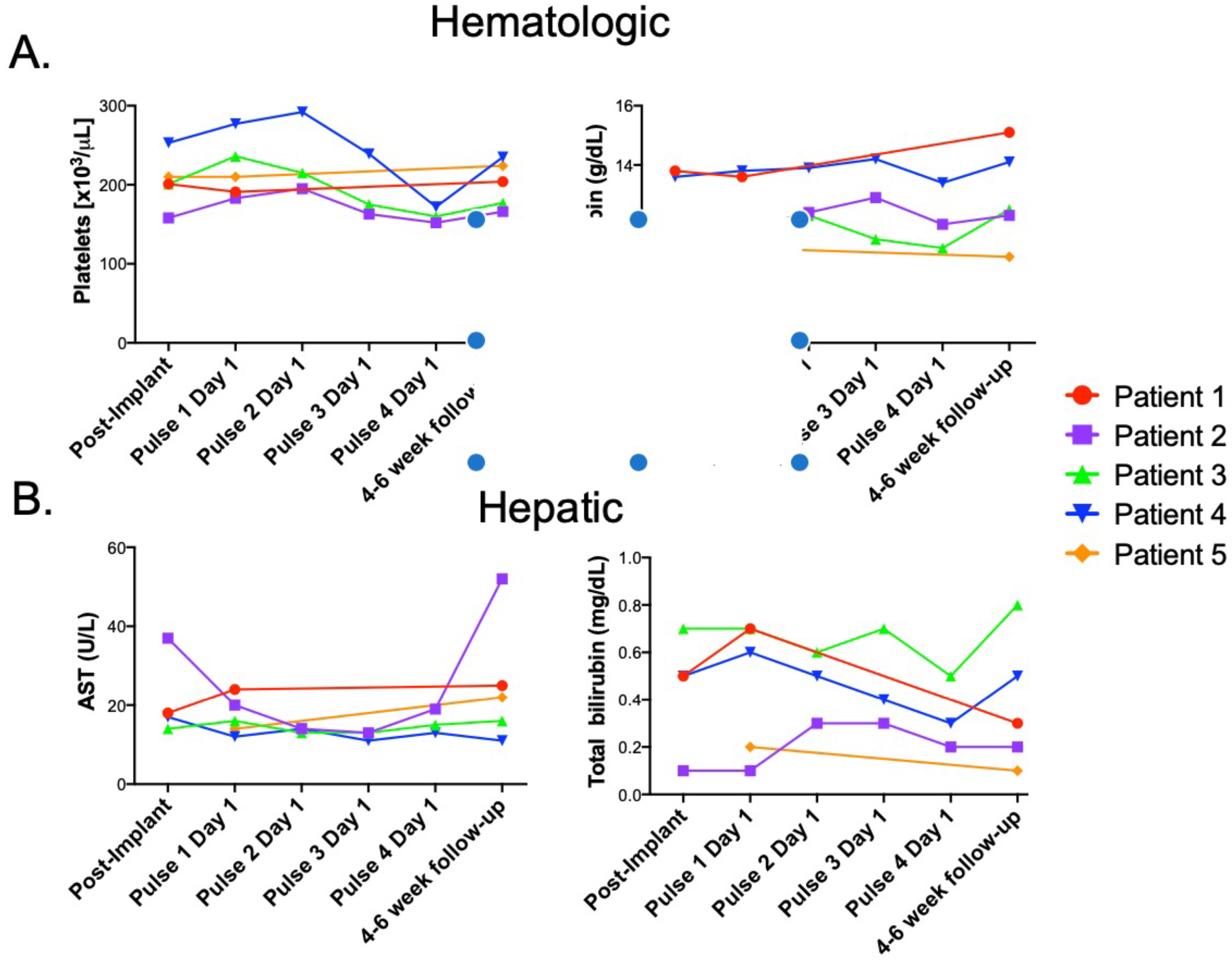
**A-B:** Laboratory values throughout the 4 pulses for platelets, hemoglobin, AST and total bilirubin to demonstrate systemic hematologic and hepatic safety.

**Supplementary Figure S3.**
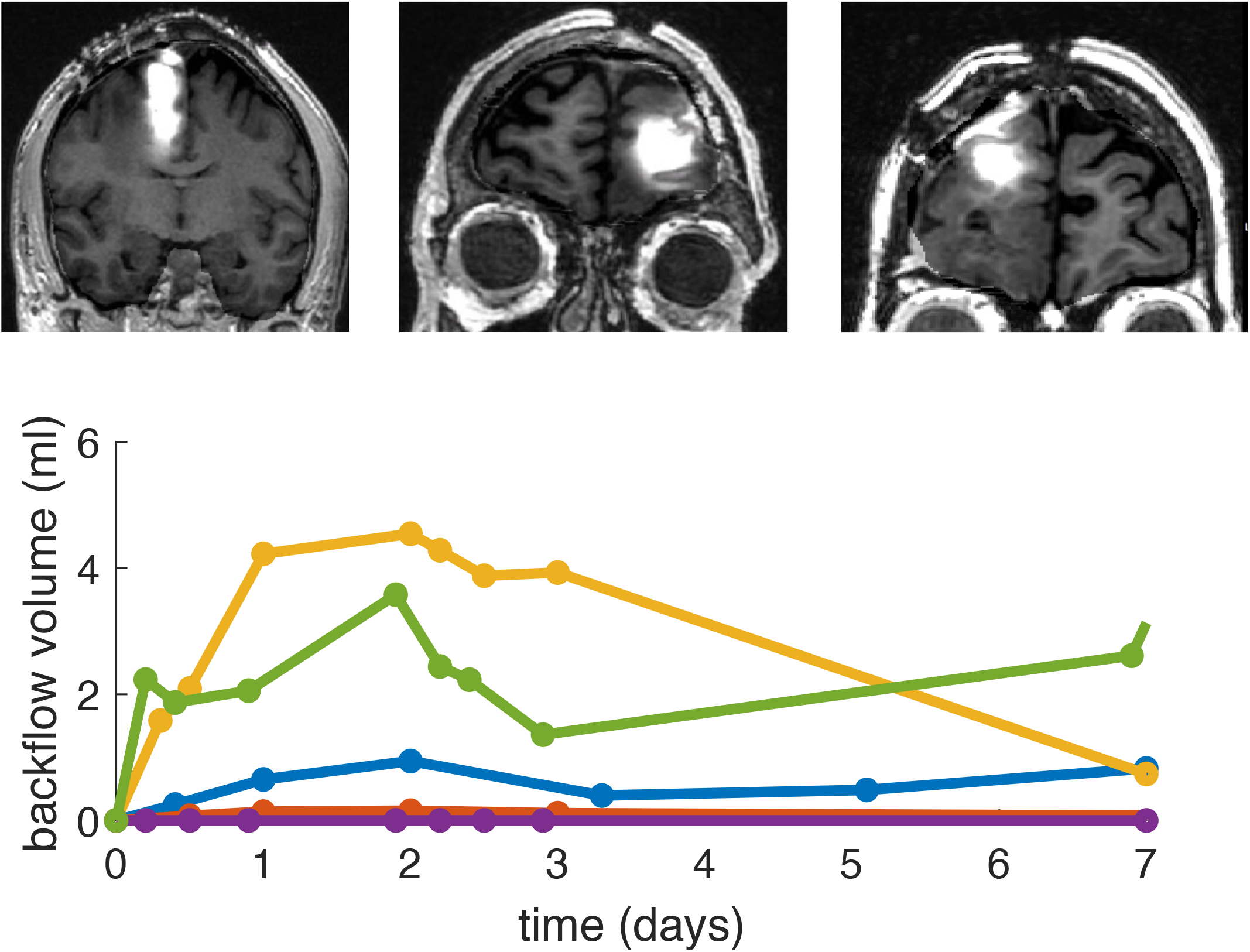
A small volume of contrast flowed back along the catheter to the brain surface (white triangles), with a time course similar to the volume infused into the brain. The maximum backflow volume was 8.8% (1.8 mL) of the volume infused into the brain suggesting that most of the drug was successfully delivered to the targeted tissue.

**Supplementary Figure S4.**
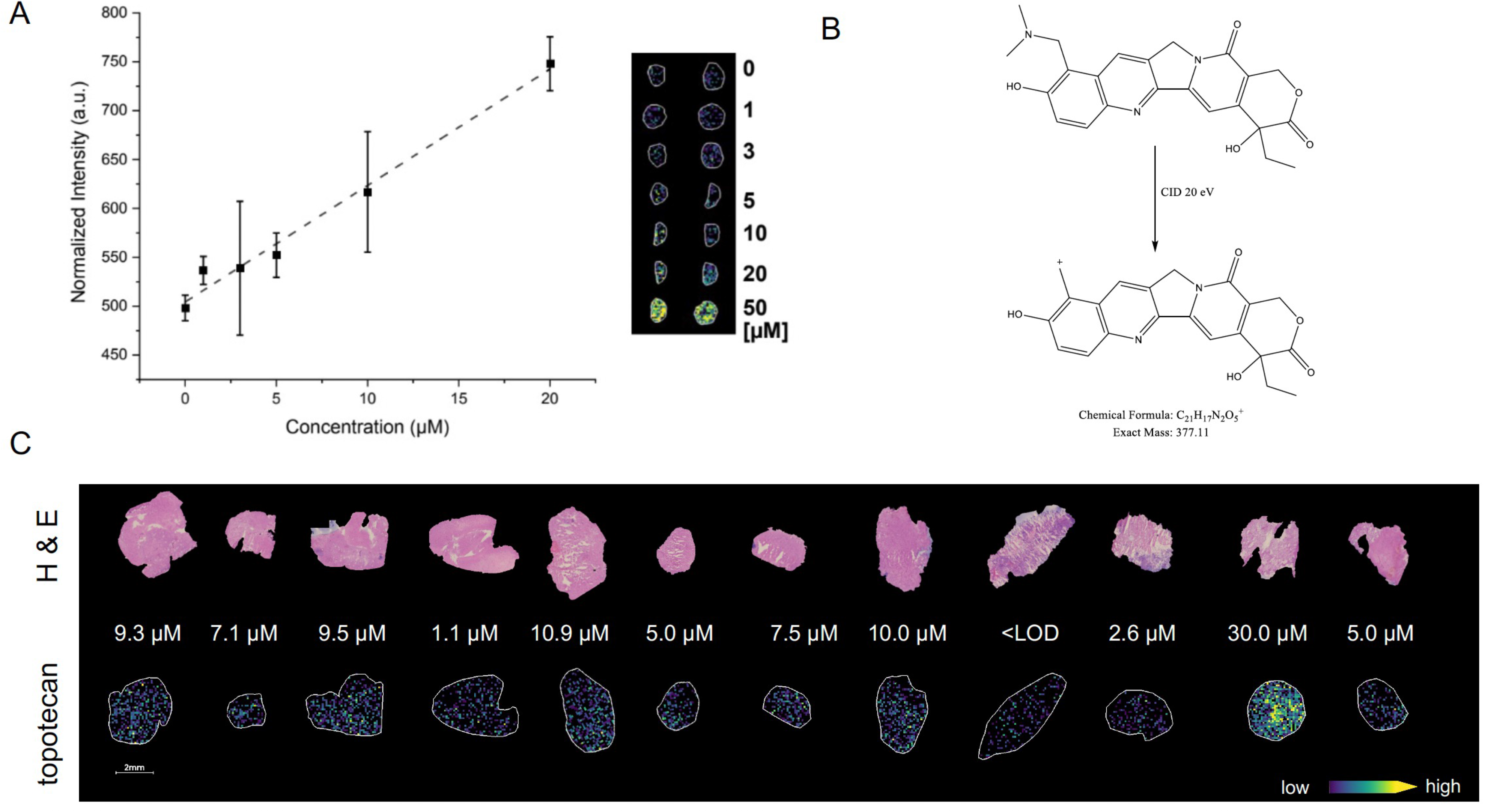
**A:** Calibration curve for TPT MALDI MSI quantification with an R^2^ = 0.984. **B:** The chemical structure of TPT precursor and product ion used for tissue quantification. **C:** H&E and TPT ion image of 12 post-CED biopsies, accompanied by estimates of TPT concentration for each sample.

**Supplementary Figure S5.**
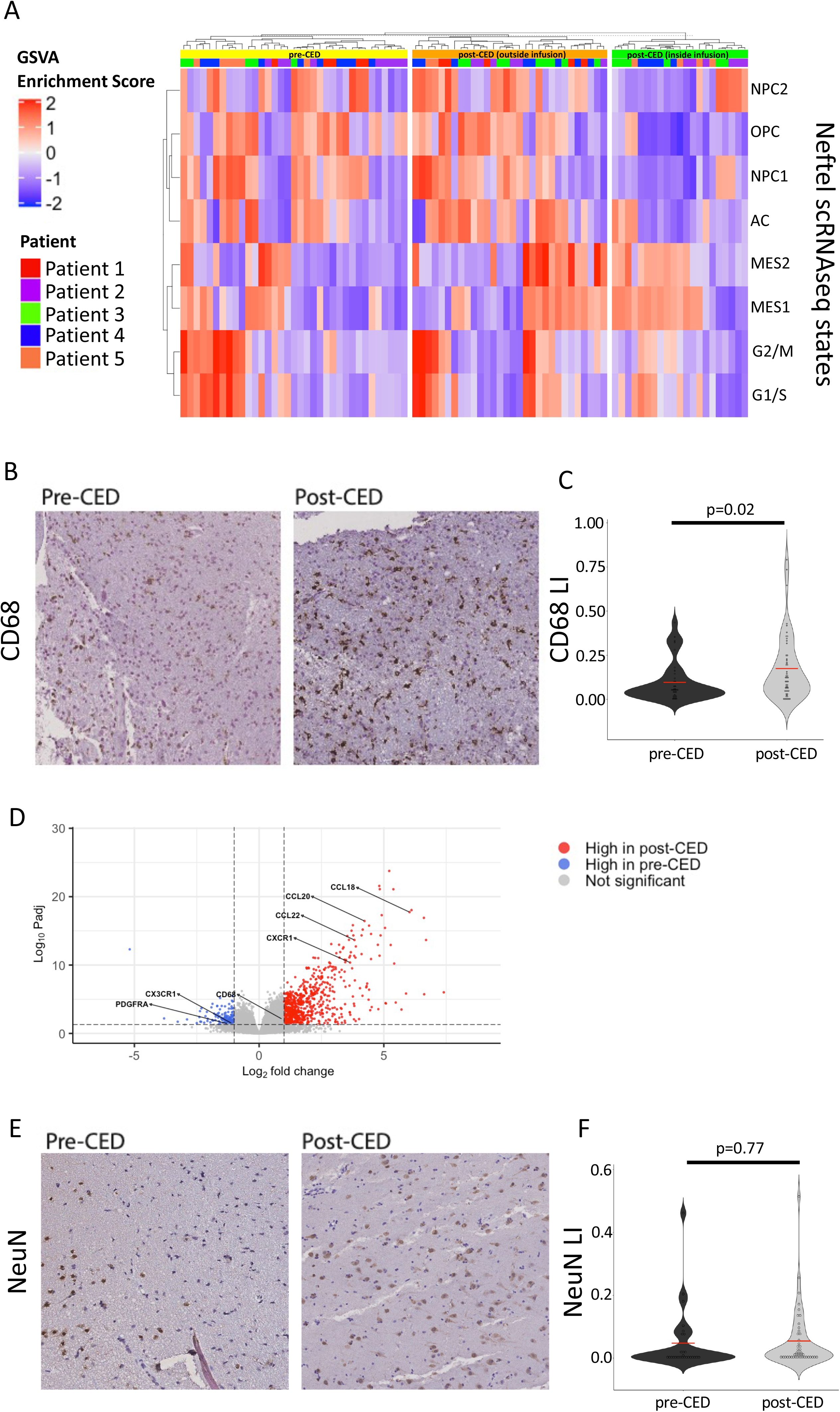
**A**: Heatmap displaying GSVA analysis of MRI-localized biopsies for Neftel et al scRNAseq-derived glioma state signatures. GSVA scores for a given gene set are scaled across all samples. On the top of the heatmap, each biopsy is annotated by patient, as well as whether the biopsy was taken pre-CED or post-CED. The post-CED biopsies were further radiographically stratified by whether they were inside or outside the maximum volume of distribution. **B**: Representative images pre-CED and post-CED immunohistochemically stained for CD68 (activated microglial marker). **C**: Violin plot displaying quantification of CD68 by labeling index across all MRI-localized biopsies from all patients, comparing biopsies taken pre- and post-CED using a student’s T-test (n=87). **D:** Volcano plot displaying differentially expressed genes between pre-CED and post-CED MRI-localized biopsies across all 5 patients (n=86). Cutoffs were log-2-fold-change > |1| and adjusted p-value<0.05. Select significantly differentially expressed immune/inflammatory-associated genes are marked. **E**: Representative images pre-CED and post-CED immunohistochemically stained for NeuN (neuronal marker). **F**: Violin plot displaying quantification of NeuN by labeling index across all MRI-localized biopsies from all patients, comparing biopsies taken pre- and post-CED using a student’s T-test (n=87).

## Supplementary Table Information

**Supplementary Table S1:** Supplemental Clinical Information [omitted to protect patient privacy, provided upon request to corresponding author]

**Supplementary Table S2:** Differential Gene Expression Analysis Post-CED vs. Pre-CED

**Supplementary Table S3:** Gene Set Enrichment Analysis Post-CED vs. Pre-CED

**Supplementary Table S4**: Glioma Tumor Cell State Classification

**Supplementary Table S5:** Differential Gene Expression Analysis Post-CED within treatment volume vs. Pre-CED

***\*Additional Safety Data Unrelated to CED Provided upon Request to the Corresponding Author\****

## Footnotes

a Detailed pathology descriptions, as well as other clinical data, are provided in **Supplementary Table S1 which is available upon request to the corresponding author.**

